# Research agenda setting in the mental health of neurodivergent students: A qualitative exploration of student’s perspectives

**DOI:** 10.64898/2026.07.29.26359210

**Authors:** Lea Satala, Daria Melashenko, Annabella Feeny, Dejla Hoxha, Sumeiyah Koya, Clara Sanchez-Izquierdo Lozano, Zepeng Long, Amanda Russell, Aja Murray, Luke Power

**Author notes:** These authors contributed equally to this work (Lea Satala, Daria Melashenko). Corresponding author at: 7 George Square, University of Edinburgh, EH8 9JZ.

## Abstract

**Objectives:** To identify research priorities for improving the mental health of neurodivergent higher education (HE) students by exploring the perspectives of individuals with lived experience.

**Design:** Qualitative study using an online survey. Data was analysed using a deductive–inductive, hybrid semantic thematic analysis.

**Setting:** UK higher education institutions.

**Participants:** 104 current and former neurodivergent HE students with diverse neurodivergent profiles and intersecting identities.

**Main outcome measures:** Participant recommendations regarding priorities for future research on neurodivergent student mental health.

**Results:** Six themes were identified and were grouped into (1) general recommendations for research and (2) recommendations specific to neurodivergence within a HE context. Participants prioritised a shift away from medical model approaches towards research informed by social and strengths-based perspectives. Key priorities included improving understanding of diagnostic barriers and misdiagnosis, reducing stigma, investigating institutional barriers within HE, evaluating the effectiveness of support and accommodations and examining the experiences of underrepresented and intersectional groups. Participants emphasised the need for research on more flexible teaching practices, sensory-friendly learning environments, integrated mental health and educational support and alternatives to diagnosis-dependent access to services.

**Conclusions:** Future research should move beyond descriptive accounts towards evaluating interventions and current support provision to understand if they improve the mental health of neurodivergent students. Adopting intersectional approaches, moving beyond binary deficit- or strengths-based frameworks and focusing on inclusive, needs-based support rather than diagnosis-led systems are likely to produce more equitable and effective outcomes for neurodivergent students in higher education.

**What is already known on this topic:**

- Neurodivergent higher education students experience poorer mental health than their neurotypical peers and face substantial barriers to accessing appropriate support.
- Although research priorities have been identified for neurodiversity in higher education, the mental health research priorities of neurodivergent students have received little attention.

**What this study adds:**

- Neurodivergent students prioritised research that moves beyond medical-model approaches towards understanding institutional barriers, stigma, diagnostic pathways and the effectiveness of support in higher education.
- Participants also highlighted the need for greater attention to underrepresented groups and intersectional experiences within research.

**How this study might affect research, practice or policy:**

- These findings provide a lived experience-informed research agenda to guide future research, funding priorities and support for neurodivergent students in higher education.

---

Approximately 15-20% of higher education (HE) students are neurodivergent [1]. Their attrition rates exceed those of neurotypical peers and poor mental health is considered a key contributing factor [2–4]. The transition to university may intensify challenges for neurodivergent individuals, as neurotypical norms dominate social and academic environments, leading to a sense of exclusion [5,6]. Fears of being labelled or misunderstood by staff and peers often result in masking behaviours, which have been associated with increased psychological distress and burnout [2,7–9]. High rates of anxiety and depression are well documented among neurodivergent students, [6,9–13]. However, mental health services often remain difficult to access [14], and the mental health burden can be further intensified for those whose neurodivergence intersects with other marginalised identities based on ethnicity, gender, or socioeconomic status [15]. Despite the urgency of these challenges, there remain critical evidence gaps to underpin better support for neurodivergent students’ mental health.

There has been growing recognition of the need for priority-setting exercises in neurodivergent mental health to guide research, as a means for ultimately informing and enhancing the impact of policies and interventions [16]. Understanding the experiences and needs of neurodivergent students from their own viewpoints is crucial for developing effective and targeted support systems and policies [17]. However, many initiatives lack meaningful involvement from autistic and other neurodivergent individuals themselves, particularly in the UK, where research funding and research aims often misalign with or misjudge the priorities of the autism community [18,19]. This disconnect is also evident in HE, where few studies actively incorporate the voices of neurodivergent students in conveying their perspectives, needs, and priorities through co-creation and user-led research approaches [17,20,21]. Moreover, while some studies have begun exploring mental health in neurodivergent student populations more generally [22,23] and exploring research priorities of the wider autistic community [24], there remains gaps pertaining to the perspectives of neurodivergent students.

Recent studies have made advances in addressing this gap. Ostaszewska et al. identified community-led research priorities through a three-stage participatory priority-setting process, exposing systematic barriers across services available to the neurodivergent community, including intersectional stigma and institutional discrimination [25]. Their findings emphasised the need for improved support across the education system. Le Cunff et al. engaged neurodivergent students, staff, and graduates to identify key priorities for future research into neurodiversity within HI [26]. They identified 10 priorities that broadly prioritise research into staff knowledge and attitudes; types of assessments that help neurodivergent students thrive; what support do neurodivergent students want; what factors impact neurodivergent students’ outcomes and accessibility of university. While these mark critical progress in aligning research agendas with stakeholder needs, more work is needed to further our understanding of research priorities around the mental health of neurodivergent students.

Our approach is grounded in the understanding that lived experience offers an integral form of expertise to our research processes. In conducting this research, we aim not only to contribute to growing literature in the areas of neurodiversity, mental health, and HE, but also to identify opportunities for the development of more inclusive and responsive systems and policies within Scotland’s HE landscape.

## METHODS

### Positionality Statement

This research was led by the Scottish Student Mental Health Research Network (ScotSMART) which brings together researchers, students, mental health professionals, and individuals with lived experience of mental health challenges and neurodivergence. We recognise that student mental health is inherently shaped by social and institutional contexts, including experiences of inequality, marginalisation, and exclusion, and that research must be co-produced with those most impacted by the systems and issues we study. As a network we acknowledge the power dynamics present in research and our varied positionalities within our research team. We aim to dismantle these power dynamics through interdisciplinary collaboration and shared ownership by individuals with lived experience.

### Participants

Participants were drawn from a larger survey about research priorities in relation neurodivergent student mental health and were those who identified as current or former neurodivergent HE students (n=104). The sample was majority females (n=73, males=14, non-binary/third gender=14, prefer not to say =2, self-described =1), with an age range of 18 - 63 (M=32, SD=9.68). Half (n=52) reported intersecting identities such as being a researcher, HE student or support worker (see Table S1 in Supplementary Materials). The majority of participants identified as having ADHD (n=18), being autistic (n=17) or both (n=15), with a significant proportion of participants reporting belonging to more than one neurodivergent group (see Table S2 in Supplementary Materials).

### Procedure

Participants were recruited through convenience sampling via social media advertisements, professional mailing lists, and researchers’ networks. They completed an online qualitative survey comprising 10 questions which asked them to consider the state of neurodivergent student mental health research. These questions derived from existing literature and iterative dialogue within the ScotSMART team. The survey also included questions exploring research priorities specific to the Scottish higher education context. Although all participants were presented with these questions, only those with experience of higher education in Scotland provided responses. The full survey is provided in Supplementary Materials. The study was approved by the University of Edinburgh Ethics Committee (246-2324/1) and all participants provided informed consent before their involvement.

### Analysis

To ensure coding consistency, members of the qualitative analysis team (DM, LS, ZL, YB, AR) independently coded a sample of the data (three transcripts) and discussed similarities and differences in their interpretations. This allowed for the development of common coding practices for consistency. We used a deductive-inductive hybrid, semantic thematic analysis (TA). This approach is considered semantic as we are interested in participant’s explicit recommendations rather than uncovering latent or implicit meaning [27,28]. In addition, it is deductive/ inductive as we used a 2-phased analytical approach (see also [29–31] where we (1) extracted data through an *apriori* template/ guide in the form of a quasi-codebook [32] and (2) searched for themes/sub-themes within this framework inductively, this included the establishment of higher order themes (see below).

In a first stage, a deductive approach involved initial exploration using a 3-code guide. This guide was utilised to organise the initial data extraction around the research questions and study aims. These three codes included: ‘knowledge of extant research’ (views on current research), ‘research required’ (the types of research participants suggest is needed) and ‘neurodivergence in Scotland’ (findings related to the Scottish context). The use of predetermined codes enabled the acquisition of data within specific, study parameters [33].

The 104 participant transcripts were distributed among 5 researchers (LS, ZL, DM, YB, AR). Using the guide, data was independently coded and extracted. Following this, 6 researchers worked in pairs (LS-ZL; DM-YB; AF-AR) to examine a specific code and assessed potential sub-codes. Each pair initially extracted sub-codes independently and then discussed similarities/ differences together.

The second, inductive, stage of analysis aimed to draw out themes associated with the codes and sub-codes. Two members of the research team (LS and DM) examined all data across the three initial codes and their related sub-codes to develop higher level themes This was an iterative process where each researcher analysed half of the data and then provided a secondary review of the other half. Any disagreement was discussed and a third researcher (LP) examined and validated the sub-themes.

## RESULTS

Six themes were generated from the analysis, relating to (a) general recommendations for research and (b) recommendations for research around Neurodivergence (ND) within a HE (HE) context.

### 1. Recommendations for Research: General

Theme 1: Shift away from research that utilises a medical model and towards one which accounts for context

Participants commented on the philosophical underpinning of research and the dominant paradigm within neurodivergent research. Multiple participants commented on the ‘medical model’ dominance, e.g., “*the vast majority [of research] works in the Medical Model*” which, according to this participant, is *“treating our brains as something that is malfunctioning and tries to fix it*.”. Further expanding on this point, they suggested that research needs to “*change to the Social Model and focus on the barriers we face and the trauma caused by those barriers*”. The need to move beyond the dominant medical model in research, was further highlighted when another participant commented on the “pathologizing” nature of the medical model:

> *“raises the question of whether what we label as “disability” is actually a mismatch between the individual’s cognitive style and societal expectations, rather than an inherent deficit”.*

Within the above quotation, the participant is reflecting on and examining the expectations which neurodivergent students experience within neurotypical, HE environments. They are critically examining what is meant by ‘disability’, specifically the way in which (dis)ability is socially mediated through cultural norms. In addition, the participant points to the deficit orientation of the medical model and the consequential construction of neurodiversity as ‘less than’. Participants noted the importance of moving beyond this approach and focusing on the barriers neurodivergent students face, including their ramifications. Thus, according to the participants, there is a need for research to adopt insights reflective of a social understanding of (dis)ability.

Participants also suggested steps that could be taken to decrease the influence of the medical model within research. For instance, one participant recommended a research approach that saw students with neurodiversity “*as a whole person, not siloed in a diagnosis*”, highlighting the compartmentalizing effect of such a model. Related to this is the need for more dedicated research using a strengths-based approach when examining neurodivergence within HE. Overcoming the deficit-based model, emblematic of the medical model, is of paramount importance given that:

> *“certain neurodivergent traits can be an asset in higher education and how to capitalise on our strengths”*

Another participant pointed to the need to capture the nuance of individual neurodivergent experiences by focusing on both the strengths and complexities that neurodiversity entails:

> *“There’s a move to a strengths-based approach which I appreciate, however I feel like the research is very much all or nothing”*

This highlights the need for research to focus on and further understand the strengths of those who are neurodivergent, specifically in an HE environment, as currently students are “*tolerated, not celebrated*”. However, this must also be balanced with a research focus that addresses the relationship between strengths and need.

Participants also commented on areas of research that reflect a reorientation towards the social model of (dis)ability, these would focus on:

> *“how […] schools and universities [can] create and implement personalized support systems that cater to the unique mental health needs of neurodivergent students, considering the wide range of conditions”.*

The above points to the types of research that should be prioritised to overcome the dominance of the medical model. Rather than seeing a student’s outcomes as a consequence of individual factors, the recommendations above highlight the need to examine the environment within which the individual is situated and how this could be tailored to the specific needs of those who are neurodivergent. An approach like this articulates one’s context as the disabling factor.

#### Theme 2: Difficulty with the diagnosis process and the issues of misdiagnosis

Another proposed area for research is the examination of the diagnosis process and the consequential impact of misdiagnosis. Very often, to receive the necessary support and accommodation, an official diagnosis is required as there is:

> *“No support in place for anyone who’s not diagnosed [which requires individuals] having to jump through hoops first”.*

However, in order to receive this diagnosis, as articulated by one of the participants, students face multiple barriers. One such barrier is waiting lists, with one participant stating that “*diagnosis in some parts of [mid-sized UK city] is currently 5-10 years*”. In addition, and related to the above point, is the financial burden this places on those requiring a diagnosis. According to participants, due to the long National Health Service (NHS) waiting lists, the only alternative option is a privately obtained diagnosis, which can be costly. Thus, given the importance of, but barriers to, receiving a diagnosis, participants point to the need for further research examining this process and methods of overcoming barriers so students can access needed support services.

Participants also commented on additional barriers related to the diagnosis process, specifically for at-risk and vulnerable minority groups. Prominent among them is the suggestion that women rarely receive a diagnosis due to “*biases in diagnostic criteria and presentation*”. Other possible underdiagnosed groups of students, as shared by the participants, include ethnic and gender minorities, care-experienced individuals, as well as for those with “*less famous*” neurodivergent conditions. Given the above, research must begin to look at the barriers to diagnosis faced by vulnerable communities and how these may be overcome.

Another area of concern requiring further research is misdiagnosis. This is highlighted by a participant who stated that:

> *“neurodivergent individuals are going their whole lives diagnosed with mental health disorders [who are then] later in life diagnosed with neurodivegent conditions and find they feel better/function better - it’s literally costing quality of lives and actual lives.”*

The above points to the risks associated with a misdiagnosis and the need to accurately distinguish between a neurodivergent and a mental health condition. Participants also commented on how this intersects with gender, with women being specifically at risk. Given these insights, participants outline the need for research to examine the experiences of misdiagnosis, how they occur, how they interact with gender; this includes a focus on preventative measures.

Conducting more research on the students who have been diagnosed later in life was also mentioned by multiple participants in the dataset.

#### Theme 3: Stigma

The last general research recommendation relates to stigma. Participants outlined their experiences of stigma, how this related to being neurodivergent, and the ubiquity of this experience for students. Participants noted that there is often a significant lack of understanding and education on neurodivergence, with one of them sharing that students may face stigma and discrimination not only from peers and society, but also from educators. The consequences of these experiences are succinctly outlined in the below quotations:

> *“[stigma] leads to social isolation, low self-esteem, and increased anxiety or depression”,*
>
> *“[those who are neurodivergent are] at higher risk of being bullied and socially excluded, which significantly impacts their mental health and sense of belonging in the educational community”.*

It is clear that students who are neurodivergent are at increased risk of mental ill-health. This is, as multiple participants suggest, linked to their struggle to be accepted by peers due to their neurodivergent trait manifestation. In addition, participants also noted that stigma may become internalized and prevent students from engaging with needed support services. Given the increased risk of stigma being experienced by neurodivergent students, research should focus on methods and interventions that target neurodivergent stigma and the subsequent creation of educational environments that minimize mental ill-health risk.

Lastly, participants also commented upon the intersection of neurodivergence with other factors (culture and sexual orientation) in experiences of stigmatization. For instance, students from racial and ethnic minorities may face additional barriers related to “*cultural stigma, language differences, and lack of culturally competent services*” while LGBT+ students who are neurodivergent, “*may face dual stigmatization and discrimination, which can impact their mental health and access to supportive communities”*. Thus, interventions focused on stigma reduction must also examine how neurodivergent experiences intersect and are shaped by other forms of stigma. It’s important for researchers to remember that, for neurodivergent students,

> *“It is a radical act to simply exist and act the way you want to even if it is not harming anyone, like openly stimming”.*

### 2. Recommendations for Research: Neurodivergence (ND) within a higher education (HE) context

Theme 4: Research to explore key barriers experienced by people who are ND in HE (including how they can be overcome):

Participants also provided recommendations for research relating to the barriers affecting neurodivergent student’s mental health in HE settings. These barriers primarily related to institutional structures within the education system and wider environmental factors.

#### Institutional structures

Participants consistently highlighted that HE is largely designed around neurotypical norms. As one autistic participant with dyspraxia questioned:

> *“How does the neurotypical standards of teaching and learning in Higher Education affect the experience and outcomes of neurodivergent students?”*

This framing reflects a broader concern that the structure of HE privileges neurotypical ways of learning, thinking and performing. Another institutional barrier identified, intimately related to the first, is inflexibility within teaching and learning practices. One participant with Attention-Deficit/Hyperactivity Disorder (ADHD) noted that:

> *“learning is too inflexible and leads students to internalise their challenges as something being wrong with them”*

The above quotation indicates that rigid systems, emblematic of those structured around neurotypical designs, can contribute to self-blame and poorer mental health among neurodivergent students. Participants emphasised, for example, that assessments are often designed with neurotypical cognitive styles in mind, as noted by a participant with ADHD and mental health conditions:

> *“the requirement for particular methods of organization and thinking to fit into the assessment process”.*

Participants further discussed exams, coursework, and deadlines as significant sources of anxiety and distress. Exams were described as particularly anxiety-provoking, while the clustering of assignments across the academic year was linked to burnout. A student support with Fetal Alcohol Spectrum Disorder (FASD) asked:

> *“How can we best manage the spread of assignments, tasks and exams throughout the year, to avoid overwhelm and burnout? (Balance between independent learning and support).”*

Another inflexible learning practice within HE institutions, as noted by participants, is course design and delivery. Alongside assessments (research projects, presentations etc.), participants commented on lecture design, inflexible deadlines and the balance between group and individual work. Group work, in particular, was experienced as challenging, with one autistic participant with ADHD stating *“the impact of group work [is] very difficult on my mental health”*. Thus, given the mental health impact of inflexible teaching within HE, there is clearly a need for further research into alterative teaching practices, including course designs, that meet the needs of those who have neurodivergence.

Participants also raised concerns about university support systems that rely on formal diagnosis as a prerequisite for access to accommodations. In the Scottish context, this was described as particularly problematic due to long NHS waiting times- as indicated previously. As a researcher and lived experience expert with multiple neurodivergence explained:

> *“It may be more difficult to get an neurodivergent diagnosis in Scotland due to the differences in the NHS compared to the rest of the UK. There needs to be research surrounding institutional approaches to self-diagnosed neurodiverse students.”*

This highlights how diagnostic gatekeeping can exclude students who nonetheless experience significant support needs. Thus, there is a need for research that examines alterative prerequisites for accessing accommodations/ supports.

Finally, academic pressure was highlighted as a broader institutional barrier. High academic and social expectations were seen to negatively affect mental health and self-concept, as reflected in the statement by a support worker with ADHD, *“the social and academic pressures of university cause self-esteem/self-worth issues in ND students”*. Overall, participants viewed institutional barriers as significant contributors to poor mental health, emphasising the need for systemic approaches to support.

#### Environment

Participants suggested that the HE environment is largely created to fit the needs of neurotypical individuals with little adaptation to those who are neurodivergent. Participants highlight that neurodivergent students might be more sensitive to certain environmental factors, such as sounds, lighting, motion, which they are unable to control in university settings. This may consequently lead to:

> *“[a] permanent state of fatigue and feelings of always being one step away from full burnout/meltdown [which may affect] academic performance and general wellbeing”.*

Further unpacking this, some participants suggest research should pay more attention to:

> *“how people can be supported by offering different types of working environment [lighting], various areas, outdoor spaces [and] into how to best adapt current academic and social environments for neurodivergent students, focusing on preventing experiences that can lead to trauma, guilt, and shame—emotional burdens that are often difficult to overcome”.*

Given the above, the suggestion could be made that further research is needed into the different ways in which HE institutions can adapt their environments to better suit the sensory needs of those who have neurodivergence.

##### Theme 5: What is currently effective

Participants also commented on the need to understand what supports/ services work, the mere presence of support within HE does not guarantee its effectiveness. The range of support, broadly termed adjustments and accommodations, was in itself considered a “*grey area*” by participants, especially in relation to what constitutes reasonable requests for support. Participants highlighted the need for more research evaluating which adjustments and accommodations genuinely benefit neurodivergent students, as reflected in the call to examine the *“effectiveness of support offered by HEIs [higher education institutions]”*. In addition, participants emphasised the need to assess:

> *“What specific interventions are most effective for different types of neurodivergence in improving mental health outcomes?”*

Further, given the diversity of neurodivergent lived experience, participants stressed that a one-size-fits-all approach is unlikely to be effective. Instead, a targeted and differentiated approach to support was seen as essential. Alongside tailored supports, participants also noted the need for further research into the ways in which support services can be developed/implemented that meet both the learning and mental health need of students with neurodivergence. This was captured by a participant with a tic disorder, who emphasised the importance of understanding:

> *“how can educational institutions and mental health services be effectively integrated to provide comprehensive, tailored support that addresses the unique needs of neurodivergent students?”*

Participants thus indicate the need for further research into (1) the effectiveness of supports and interventions and (2) the ability of services to deliver integrated systems that address both learning and mental health needs.

##### Theme 6: Need for greater examination of experiences of underrepresented communities and how these experiences intersect

Participants emphasised the need for greater research attention on excluded and marginalised groups, with one participant noting that “*any groups that aren’t white men need more research attention*”. These underrepresented experiences clustered across four intersecting domains: socio-demographics, neurodivergence type, student types, and lived experiences.

Socio-demographic characteristics included gender, specifically women, Non-Binary and Transgender Individuals; age e.g., neurodivergent older people; sexuality, including those within the LGBTQIA+ community, and ethnicity Pertaining to underrepresented types of neurodivergence, participants stressed the importance of research investigating those who (1) have multiple diagnoses e.g., “*2+ diagnoses such as AuDHD*”, (2) have a co-occurring disability and (3) are non-speaking/ minimally verbal neurodivergent. Student groups identified as requiring greater research attention included mature students, postgraduate students, international students, and first-generation students. Lastly, and related to the above, participants noted the importance of research examining how the experiences of being a student and having neurodivergence intersects with socio-economic disadvantage, care experience, migrant or refugee backgrounds, experiences of homelessness and/or incarceration and having English as a second language. Given the above recommendations, it is important that future research examines the intersecting experiences of neurodivergent students from underrepresented communities.

## DISCUSSION

A central finding of this study is the need for research to move beyond a predominantly medical model of (dis)ability. The problematization of the medical model’s dominance within neurodivergent research and research into disability generally, is not unfamiliar. The idea of neurodiversity is an attempt to reclaim the research landscape from this dominant paradigm [34]. These models do not fully abandon the biological dimensions of neurodiversity but present a holistic approach which looks at the interaction between biological and social mechanisms. However, there has been consistent issues with their application and the medical understanding of (dis)ability is still prevalent [35].

Importantly, our findings suggest that this proposed theoretical shift has implications for practice, particularly in relation to diagnostic systems. Participants highlighted significant challenges associated with diagnosis, including long waiting times and barriers to access. For example, some young people in the UK wait over 2.5 years for an autism diagnosis [36). Such delays reflect the limitations of a system that positions diagnosis as the gateway to support.

Stigma emerged as a closely related theme, particularly in relation to ADHD and autism [37,38]. Focusing specifically on students who are autistic, Underhill et al., (2024) found that students with a diagnosis typically avoid stigma by (1) hiding trait-related behaviours, (2) engaging in behaviours deemed socially valorised and (3) not disclosing a diagnosis to peers. Syharat et al., found that stigmatizing labels associated with an individual’s diagnosis led to students engaging in masking behaviours to hide neurodivergent traits [39] (see also [40]). This, according to the authors, led to increased cognitive-emotional load, mental health struggles and burnout, highlighting the implications of high-stigma educational environments on student’s mental health. However, Stanek and Mattson found that students within high privilege education environments were more likely to disclose their diagnosis because stigma was low, meaning that disclosure was not seen as damaging to their social capital [41]. The impacts of stigma on those with varying types of neurodiversity, including multiple diagnoses were also highlighted by participants. Some previous work has highlighted that stigma varies by condition [42]; however, there is currently minimal research examining the stigma experienced by those who have multiple diagnoses, especially in HE.

Participants also commented on the need for research to examine the ways in which neurodivergence intersects with other identities. Bonnette et al. has suggested that, within quantitative research, there should be an ‘intersection variable’ that accounts for the ways in which (dis)ability intersects with other identities/inequalities to shape student experiences within HE [43]. Bayeh and Ryder found higher rates of neurodivergent students with certain intersecting identities and pointed to the fact that these intersecting identities shaped their HE experiences and led to greater mental health difficulties [44].

Our findings also point to a need for research examining how diagnostic processes and access to support can be improved for neurominoritized communities and vulnerable groups. Ethnic and gender minority groups were identified as being at heightened risk of underdiagnosis, despite evidence suggesting they are more likely to identify as neurodivergent [45]. Care-experienced individuals were also highlighted as an under-recognised group facing barriers to diagnosis, consistent with research on unmet mental health needs in this population [46]. Participants also drew attention to the relative invisibility of less widely recognised neurodivergent conditions, such as dyscalculia and dyspraxia.

Participants also emphasised the need for research on the role of institutional environments in shaping student outcomes. In particular, they identified a need to move beyond individual-level adjustments and understand, through further research, inflexibility within HE across teaching and learning practices which privilege neurotypical modes of learning and performance. We also identified the need for research on how universities can implement sensory adaptations to accommodate sound, lighting and motion sensitivities. Persistent gaps in this area have been documented, with interventions such as quiet spaces and sensory-friendly libraries shown to have positive effects [47].

Academic pressure was also identified as a significant concern deserving further research. While this aligns with broader student mental health literature [22], our findings highlight how such pressures are experienced in distinct ways by neurodivergent students, often exacerbating feelings of shame and reinforcing mismatches between institutional expectations and individual needs.

Finally, our findings point to a critical gap in understanding what forms of support are actually effective in HE contexts. Previous qualitative research has shown that students with disabilities often find accommodations ineffective [48]. In response, a broader adoption of an intervention science framework to neurodiversity in HE research is needed: rather than simply documenting the presence of support, future work must examine what works, for whom and under what conditions [49]. This must also address the narrow focus of existing intervention research, with many neurodivergent conditions remaining entirely overlooked. Ross et al. (p. 9), for example, found no interventions specifically targeting HE students with “dyscalculia, dysgraphia, dyspraxia […] or Tourette’s” in their review [50].

### Strengths and Limitations

The study benefits from a large and diverse qualitative sample (n = 104) from across the UK and captures a wide range of neurodivergent profiles and stakeholder roles. It also includes groups often overlooked in neurodiversity research, such as women, gender-diverse individuals, and older participants. The study integrated lived experience across all stages of the research process. The questionnaires were co-produced with PPIE groups, and key members of the research team had neurodivergent conditions themselves.

Data was, however, collected via an online qualitative survey. While this was intended to enhance accessibility for neurodivergent participants, it limited opportunities for real-time follow-up questioning from researchers and clarification from participants. Further, although the survey was co-produced, one participant noted that it did not clearly communicate that prior research knowledge or opinions were expected, and some respondents expressed uncertainty about research or areas requiring further investigation. Finally, the sample was self-selecting and recruited through academic, professional, and community networks, as well as social media. This may have resulted in a sample skewed towards more engaged or reflective individuals, potentially underrepresenting those who are less connected to such networks or more marginalised within HE.

## CONCLUSION

Future research should move beyond predominantly descriptive work and focus on evaluating existing interventions and support systems. It should examine environmental adaptations and how sensory modifications and more flexible learning environments influence wellbeing, cognitive load, and academic outcomes for neurodivergent students. Our findings also highlight the need to reconsider the current reliance on diagnosis-led systems. Future work should explore alternative, needs-based approaches to assessment and support. Consistent with participants’ emphasis on underrepresented groups, future research should prioritise intersectional approaches. Finally, future research should move towards more balanced conceptualisations of neurodivergence, recognising both strengths and challenges. This includes moving beyond binary deficit- or strengths-based frameworks towards more nuanced accounts that reflect the complexity of neurodivergent lived experience that entail both.

## Data Availability

The qualitative data generated and analysed during the current study are not publicly available because participants did not provide consent for public data sharing.

## ACKNOWLEDGEMENTS

We would like to thank all participants for generously sharing their experiences, which made this research possible.

We are grateful to Alex Bowen, Shufang Cai and Linda Collins for their help with the project. We also thank Yuti Bang for her valuable contribution to the first and second phases of thematic analysis.

We would like to acknowledge the Edinburgh Student Support Team, particularly Laura Gaine, for sharing insights into university student support structures.

